# The effect of singers’ masks on the impulse dispersion of aerosols during singing

**DOI:** 10.1101/2021.07.20.21260853

**Authors:** Matthias Echternach, Laila Herrmann, Sophia Gantner, Bogac Tur, Gregor Peters, Caroline Westphalen, Tobias Benthaus, Marie Köberlein, Liudmila Kuranova, Michael Döllinger, Stefan Kniesburges

## Abstract

**Background:** During the Covid-19 pandemic, singing activities were restricted due to several super-spreading events that have been observed during rehearsals and vocal performances. However, it has not been clarified how the aerosol dispersion, which has been assumed to be the leading transmission factor, could be reduced by masks which are specially designed for singers.

**Material and Methods:** 12 professional singers (10 of the Bavarian Radio-Chorus and two freelancers, 7 females and 5 males) were asked to sing the melody of the ode of joy of Beethoven’s 9^th^ symphony “Freude schöner Götterfunken, Tochter aus Elysium” in D-major without masks and afterwards with five different singers’ masks, all distinctive in their material and proportions. Every task was conducted after inhaling the basic liquid from an e-cigarette. The aerosol dispersion was recorded by three high-definition video cameras during and after the task. The cloud was segmented and the dispersion was analyzed for all three spatial dimensions. Further, the subjects were asked to rate the practicability of wearing the tested masks during singing activities using a questionnaire.

**Results:** Concerning the median distances of dispersion, all masks were able to decrease the impulse dispersion of the aerosols to the front. In contrast, the dispersion to the sides and to the top was increased. The evaluation revealed that most of the subjects would reject performing a concert with any of the masks.

**Conclusion:** Although, the results exhibit that the tested masks could be able to reduce the radius of aerosol expulsion for virus-laden aerosol particles, there are more improvements necessary to enable the practical implementations for professional singing.

## Introduction

During the COVID-19 pandemic, professional singing and singing in choirs or churches have been widely restricted. The reason was that there were fatal transmission events in choirs and churches, which has been assumed to be caused by singing activies^1-4^. Consequently, the person-to-person transmission pathway has been in focus of many recent scientific publications in order to understand, influence, or block parts of the transmission pathways, also for singing.

In general, it has been postulated that CoVID19 could be transmitted by direct contact or transmission of small droplets with diameters ≥ 5 μm and aerosol particles with diameters ≤ 5 μm^5-7^. The transmission pathway includes (a) the production of potentially virus-laden droplets and aerosols, (b) the impulse spreading, (c) the dispersion of aerosols by convectional flow, and (d) the inhalation of particles or aerosols by the recipient ^8^. In contrast to droplets which follow ballistic characteristics and fall to the ground within approximately 1.5 m^9,10^, aerosols could remain in the air over a long time and could accumulate, especially in closed rooms^11-13^.

The reason that singing was restricted is associated with current research findings that singing and speaking produce a much higher amount of aerosol in comparison to breathing ^7,14,15^ In this respect, it has been shown that the emission rate of aerosol particles is about three times higher for speaking and 10 times higher for singing in comparison to breathing^14^. In one case, Mürbe et al. showed a 100 times higher aerosol generation for singing in comparison to speaking^7^. Since the aerosol generation is associated with the voice production itself, a blockage of this column of the transmission pathway appears problematic if singing is performed.

A reduction of the transmission risk, however, could be achieved at the next part of the transmission pathway, which is the impulse dispersion. During the impulse dispersion, a rather high concentration of aerosols is emitted into small volume in the room, before a convective distribution starts that decreases the aerosol concentration. Using Schlieren technique, Becher and colleagues found distances for singing of more than .85m^16^ Another study with inhaled artificial aerosols showed that for singing with a well pronounced text the impulse dispersion can reach maximum distances of 1.4 m^17^. Consequently, it could be assumed that aerosol concentration is lower in greater distances of 2.0 - 2.5 m to the front due to the convective distribution. For singing of lyrics, the distance of the impulse dispersion of aerosol is dominated by the articulation of pronounced consonants^18^. Singing only on vowels was associated with much shorter ranges than singing a text^17^. Although it could be assumed that safety distances could reduce the transmission risk, it should be noted that singing activities can never be completely “risk free”, because after the expulsion, the aerosols are still in the air and only their concentration is decreased by the dissolution and distribution due to convectional flows. Therefore, aeration of closed rooms still appears important^19,20^.

Another approach to block the impulse dispersion of aerosols could be the use of masks. There have been extensive studies during the CoVID19-pandemic, exhibiting that masks could inhibit aerosol spreading in breathing, speaking and also for single cases in singing^18,21-23^. However, professional singing with masks appears limited, due to the fact that surgical or FFP2 masks change acoustic characteristics^24-28^, cause a dry mouth and limit the mouth opening which is crucial for singing^29^. Due to these limitations, there have been many innovations during the pandemic concerning developments of “singers’ masks”, i. e. masks, which enable singing while reducing the associated risk of a virus transmission. However, to the best of the authors’ knowledge, the effects of singers’ masks with respect to the impulse dispersion of aerosols have not yet been analyzed.

This study investigates the impulse dispersion of five different singers’ masks in professional singers and maps their assessment of practicability for singing performances.

## Material and Methods

After approval from the local ethics committee (no. 20-1065), initially 13 professional singers were included (11 from the Bavarian Radio Chorus, Chor des Bayerischen Rundfunks, and 2 freelancers). One subject had to be excluded from the analysis because of intensive cough during the experiment. Nine of the singers had already been part of the previous study^17^. The experiments were conducted 8 months after the first experiments. By means of the medical history and the Voice Handicap Index ^30,31^there were no voice complaints raised by any of the subjects. Further, also breathing difficulties were excluded by medical history and spirometry (ZAN, Inspire Healthcare, Oberthulba, Germany).

### Tasks

Corresponding to the previous experiments^17,18^, all subjects were asked to sing the first four bars of the main melody of Beethoven’s “Ode to Joy” with the original German lyrics “Freude schöner Götterfunken, Tochter aus Elisium” in D-major, starting with F#3 (*f*_*o*_ approx. 185 Hz) for the male and F#4 (*f*_*o*_ approx. 370 Hz) for the female voices, respectively, Figure 1.

**Figure 1:**
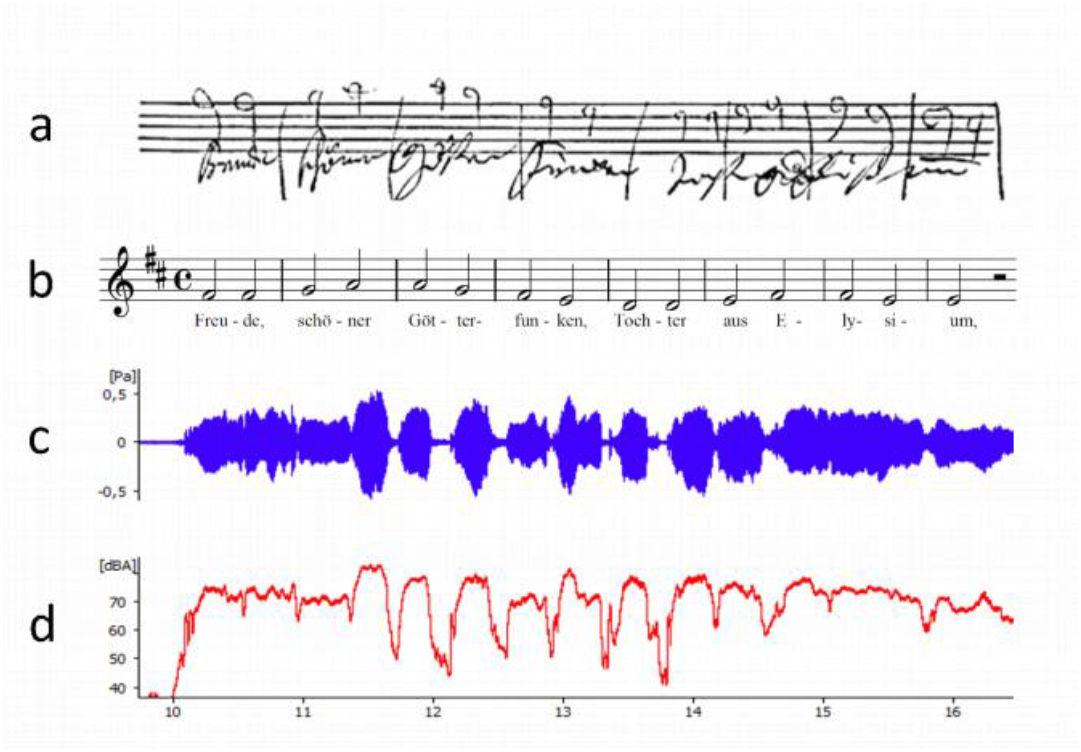
a) task in the original writing by Ludwig van Beethoven, b) task in the record, c) audio signal for a female subject, and d) sound pressure level.

The speed of approximately 80 – 90 bpm was provided by the experimenter before the experiment started. Consequently, the sung phrase had a duration of approximately 6 s. For the presented experiments, only the loud version (melody and text: MT+) of the task was performed, because the dispersion for the loud version of the sung text had shown greater values than for the soft versio^17^The end of the task was set at the end of the word “Elisium” defining the zero point in the time domain. Consequently, the start and performance of the task is represented by negative time values and the dispersion after the end of the task had positive values.

All tasks were performed immediately after the subjects inhaled 0.5 l of the vapor of an e-cigarette filled with basic liquid (50%:50% glycerin: propylene glycol). The liquid was nebulized using a Lynden Vox e-cigarette (Lynden GmbH, Dürmentingen, Germany) and the amount of gas inhaled was controlled using a ZAN 100 spirometer (ZAN, Inspire Healthcare, Oberthulba, Germany) mounted on the e-cigarette. In order to avoid secondary convective flow generated by body motions, the subjects and the investigators in close proximity were instructed to stand still for about 1min before the subject started the task. As reported by Ingebrethsen Cole et al. in 2012 the exhaled aerosols of the nebulized liquid gas have a diameter of 250 – 450 nm ^32,33^, which is about the size of respiratory aerosols transmitting the CoVID 19 virus^5-7,34^.

### Masks

In the presented study, five masks – as shown in Figure 2 – have been analyzed in comparison to singing without mask:

**Figure 2:**
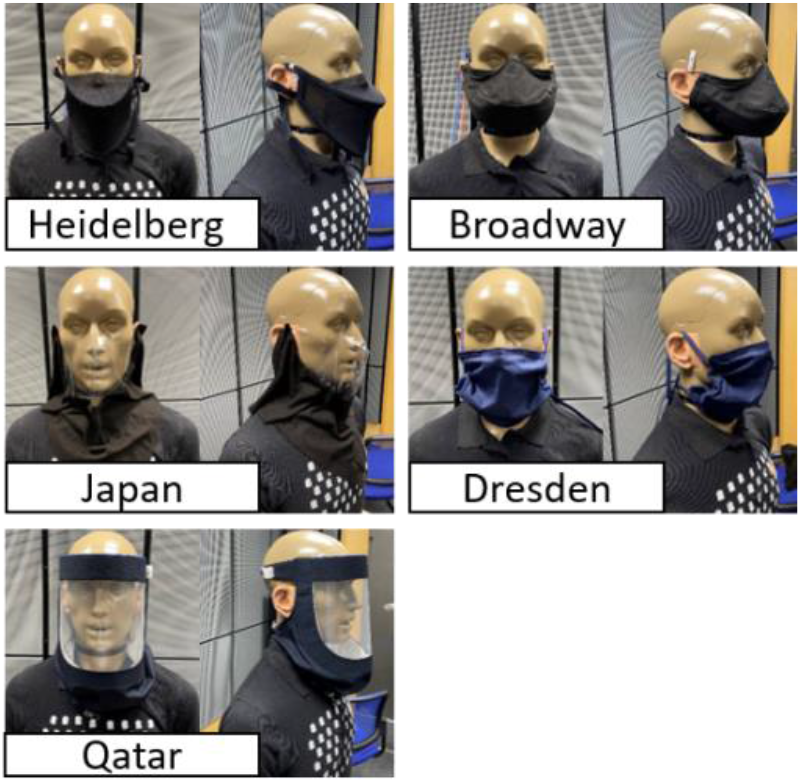
Images of all 5 masks in a front and side view perspective worn by a KEMAR head simulator.

1. The Insono-mask © (“Heidelberg Mask”); made from two layers of 6% elastane with a weight of 75g/m². The mask is fastened with two elastic straps around the head.
2. The “Broadway Mask”^35^ consists of three layers. The outside and inside layer, containing a medium weight 120 thread count cotton muslin with approximately 135g/m^2^. In between a non-woven 100% Polyester interface layer is included. This interface layer is only located in the front area of the mask. To allow an additional air supply, two small windows are cut in the fabric near each ear. Two adjustable elastic bands on each side around the ears allow to correctly wear the mask.
3. The “Japan Mask” ^36^is inspired by a design from the Nagoya Nikikai Opera, which was made to ensure their students safety while practicing. A 200g/m2 cotton molton bandana sewn to a flexible plastic face shield that only covered nose and mouth. The cotton part of the mask was knotted around the neck to cover the front part of the neck and the décolleté. The back was not sealed. The face covering can be worn around the ears. An additional elastic strap was sewn onto the plastic shield, to ensure a tight attachment.
4. The “Jazzchor-Dresden-Singmaske” (“Dresden Mask”) ^37^was invented by and for the Dresdner-Jazz-Choir. 25×60cm cotton with a weight of 255 g/m^2^ was sewn into a baggy mask to enable good mouth movement. Two ribbon-like cotton extensions on each side can be knotted tightly at the back of the head or around the ears.
5. The SannaShield mask (“Qatar Mask”): The mask was made out of a two-layers of linen cotton with a weight of approximately 120g/m^2^ which covered a typical face shield. The face shield could be mounted with an elastic band around the head and fitted on the front head. The mask can be tightly fit around the throat by tighten two cords with cord below the chin and behind the head.

All subjects tested the masks without the vapor and rated each mask on visual analogue scales of 100mm. The questionnaire contained the following questions:

1. Is the mouth opening restricted, so that singing is impaired (scale from 100 “applies very much” to 0 “does not apply at all”)?
2. Is quick inhalation for singing possible through the mask? (scale from “applies very much” to “does not apply at all”)?
3. Is the shaping of vowels/sounds limited by the mask (scale from “applies very much” to “does not apply at all”)?
4. Does dry mouth occur (scale from “applies very much” to “does not apply at all”)?
5. I can imagine singing concerts with this mask (scale from “yes” to “no”).

### Recordings

Comparable to the previous studies^17,18^, all experiments were performed in a television broadcast studio of the Bavarian Broadcasting. The size of the studio amounted 27m x 22m with a wall height of 9m. The subjects performed all tasks on a height-adjustable stage platform that was altered to fit each singer to the camera perspective. A stage mark and a mark at the forehead helped to ensure the right position for every subject. Benchmark bars in all three directional dimensions were placed on the stage. Thereby, the pixel distances of the recorded video could be translated into a metric distances. In order to reduce convectional flows, the temperature of around 21.0 °C and the humidity of around 29.8 %. were kept stable. Before each task, the studio was aerated with the main gate open for at least 2min and a fan behind the platform. After aeration, all doors were closed and the subjects were asked not to move around until all experiments were ended. Three full HD Sony HDC 1700R television cameras (Sony, Tokio, Japan, resolution 1920 × 1080 pixels) provided by the Bavarian Broadcasting service were positioned in the following way: Camera 1 (C1) recorded a side view, camera 2 (C2) a front view, camera 3 (C3) a view from above. The three cameras started recording at the same time with framerate of 25 fps. To create as much contrast as possible, the studio, stage and e-cigarette was covered in black and the singers wore black clothes and the vapor cloud was illuminated with three spotlights in a backscattering arrangement. The audio was recorded by a directional microphone (Sennheiser KMR 81), and an omnidirectional microphone ME62 Sennheiser electronic, GmbH, Wedermark, Germany).

### Video analysis

The measurements of distances reached by the vapor for all three dimensions (to the front: x-axis, to the sides: y-axis and in the height: z-axis) were extracted from the video material of the cameras 1 and 3, Figure 3. In images of camera 2, however, the contrast was reduced due to the e-cigarette equipment and light reflexes on the plexiglass shielding in the background. Consequently, these images have not been used for the detection of distances. The videos of cameras 1 and 3 were converted into a greyscale and inverted. Bright non-vapor regions as the subjects’ skin and the benchmark bars were masked in black to avoid false segmentation. The area of the exhaled vapor cloud was finally segmented for each video frame with the software Glottis Analysis Tools ^38,39^(University Erlangen-Nürnberg, Germany), using a threshold-based region-growing algorithm. The area of the cloud as well as the contour were measured as a function of time. The Region of Interest (ROI) amounted *ROI*_*x*_ × *ROI*_*z*_ = 3.8*m* × 2.9*m* for camera C1 and *ROI*_*x*_ × *ROI*_*y*_ = 2.0*m* × 2.4*m*. After the segmentation, the maximum expansion of the cloud in dx, dy, and dz. Thereby, camera 1 served as basis for the dx- and dz-, and camera 3 for the dy-expansion.

**Figure 3:**
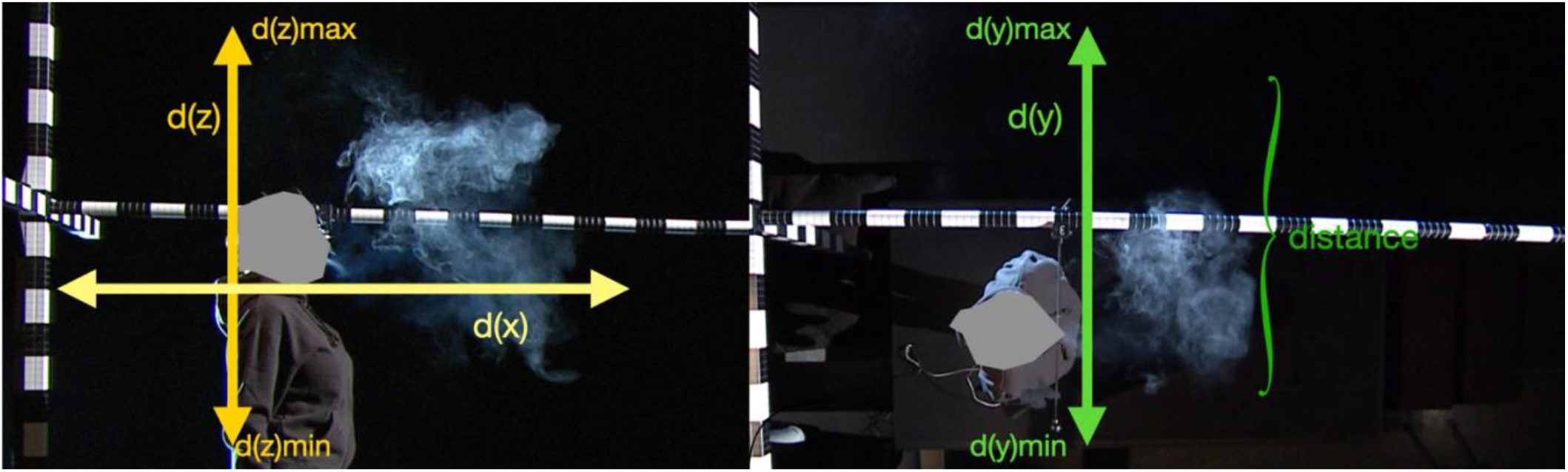
X-dimension to the front, y-dimension to the side and z-direction as extracted from camera 1 (left) and camera 3 (right).

In a final step, the resulting temporal devolutions of dx, dy and dz were filtered by a moving median filter with a window length of 30 time points and smoothed by cubic spline interpolation to remove outliers from the segmentation process, as described in detail in a previous publication^18^. This computation of curve smoothing was performed in Matlab (The Mathworks Inc., Natick, MA).

### Statistics

Due to a limited number of subjects, comparative statistical analysis appeared not meaningful.

## Results

All but one subject succeeded in performing the experiment. However, many subjects noted that singing with the vapor was associated with changes in the self-awareness of singing. Figure 4 shows the frontal and side view of a female subject for all masks and singing without mask at the end of the task.

**Figure 4:**
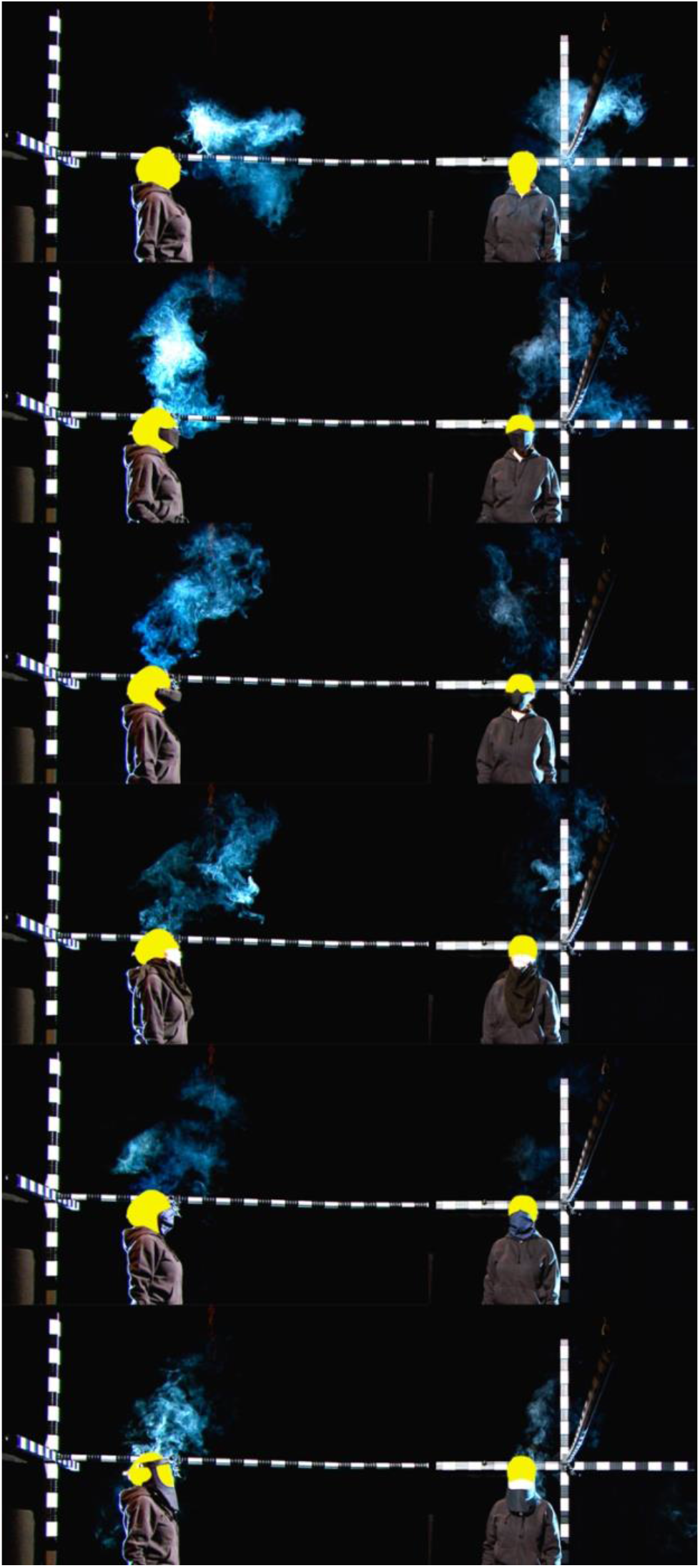
Side (camera 1, left) and frontal view (camera 2, right) of a female subject at the end of the task.

For singing without mask, the median distance of the aerosol vapor impulse dispersion to the front (dx) showed .76m at the end of the task, Figure 5.

**Figure 5:**
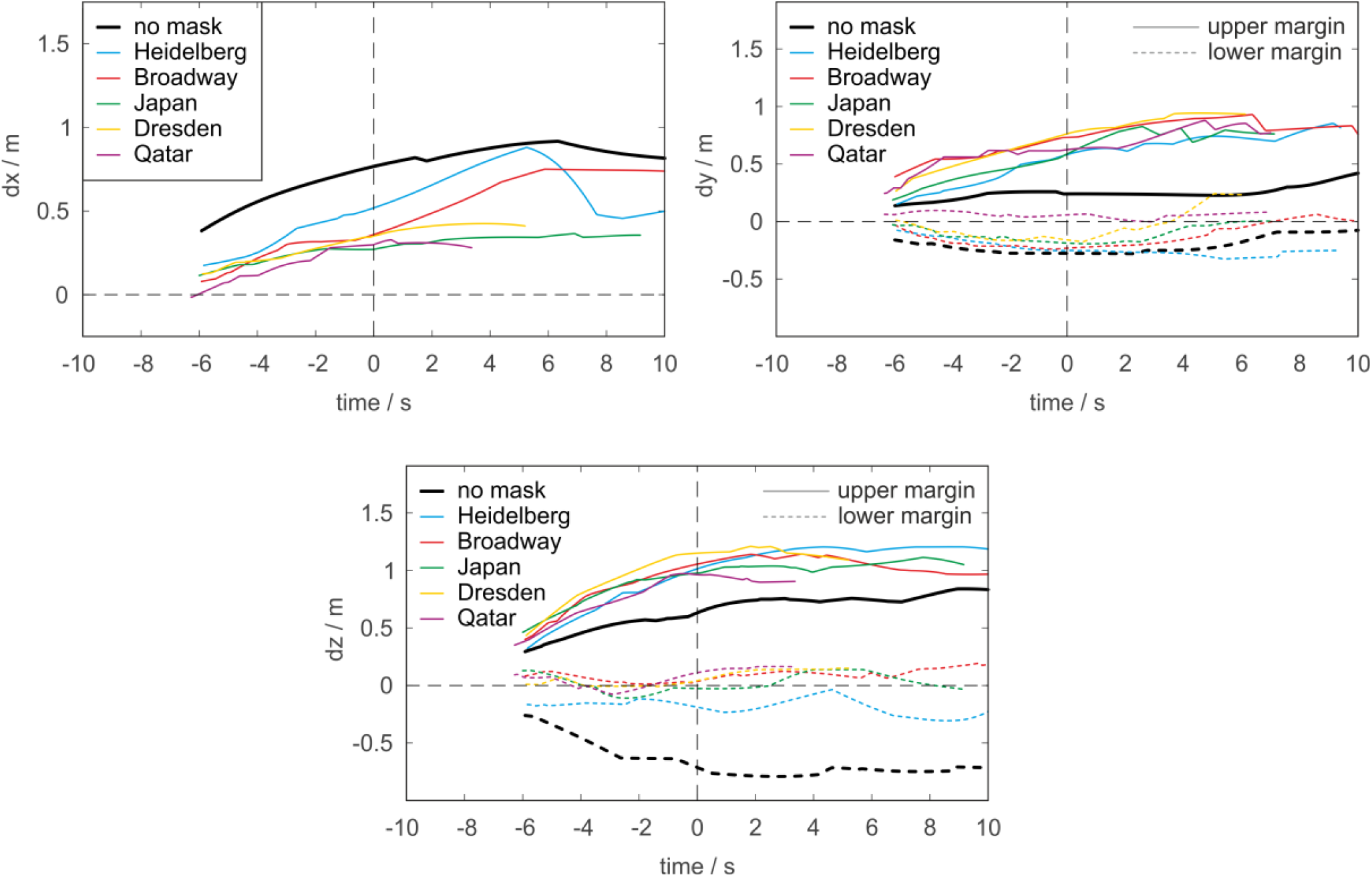
Median curves for the dispersion to the front (x-direction, upper row left), to the sides (y-direction, upper row right), and top/bottom (z-direction, lower row) for all masks and singining without any mask.

However, as can be seen in Figure 6 a single subject reached a maximum distance of 1.14m.

**Figure 6:**
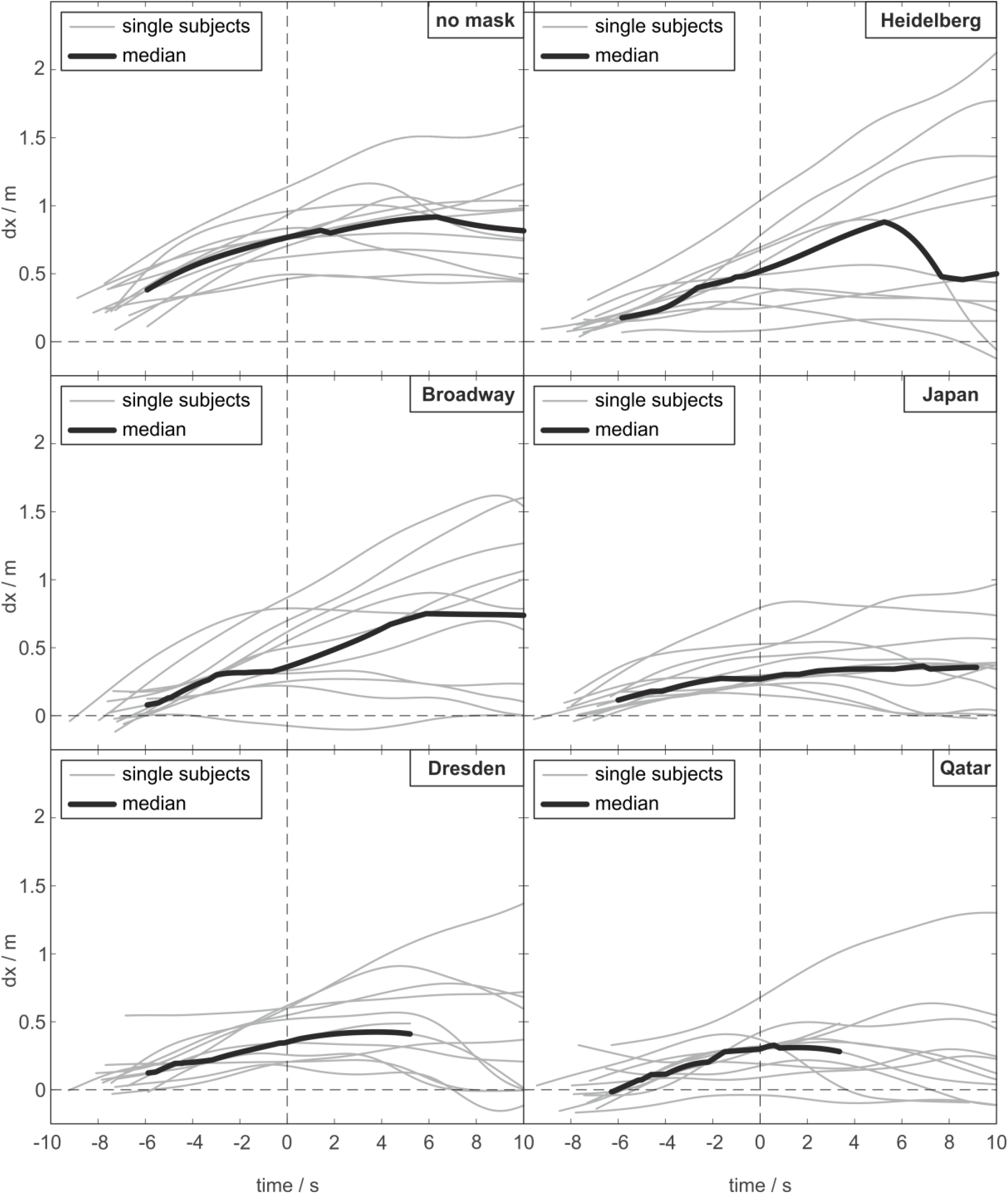
Impulse dispersion to the front for all subjects (grey) and the median (black) with regard to singing with all masks and without any mask.

The median dispersion to the side exhibited lower values with 0.25m to the left (positive dy) and -0.26m to the right (negative dy) with maximum values of 0.85m to the left and -0.69m to the right side.

The expansion of the vapor to the front was diminished by all of the masks, Figure 5. The maximum dispersion to the front at the end of the task was below 1m for all masks, Table 1. The only exception was the Heidelberg Mask, which had a maximum dispersion of 1.04m, which is still less than singing without a mask. The medians showed greater dy values for singing with the masks with respect to the dispersion to the left (positive dy values) but not to the right (negative dy values). The maximum diameter of the cloud in y-direction among all subjects was .98m without a mask, while the Heidelberg Mask amounted to 1.33m, Broadway to 1.54m, Japan to 1.19 m, Dresden to 1.20 m, and Qatar to 1.51 m. In vertical direction (dz), all masks showed a deviation of the median distances in upward direction as shown in Figure 5.

**Table 1:** Maximum and minimum values across all subjects concerning the impulse dispersion at the end of the task concerning x-direction to the front, y-direction to the sides and z-direction to the top and bottom.

| End of task (0)s | x-direction | y-direction |  | z-direction |  |
| --- | --- | --- | --- | --- | --- |
|  | Max [m] | Min[m](right) | Max[m](left) | Min[m](bottom) | Max[m](top) |
| No Mask | 1.14m | -0.69 | 0.85 | -1.29 | 1.17 |
| Heidelberg | 1.04m | -0.51 | 1.07 | -0.52 | 1.22 |
| Broadway | 0.87m | -0.94 | 1.14 | -0.18 | 1.22 |
| Japan | 0.79 | -0.32 | 1.04 | -0.70 | 1.16 |
| Dresden | 0.62 | -0.45 | 1.16 | -0.66 | 1.25 |
| Qatar | 0.68 | -0.51 | 1.14 | -0.62 | 1.11 |

All tested masks were associated with limitations concerning the singing performance without inhaling the vapor, Figure 7. In this respect, most subjects estimated the use of any of the masks as undesirable for performances on stage. Some of the masks restricted the mouth opening. With exception of the Heidelberg Mask, all other masks showed an impairment of sudden inhalation patterns. An associated dryness of the mouth due to the mask was most present for the Dresden mask.

**Figure 7:**
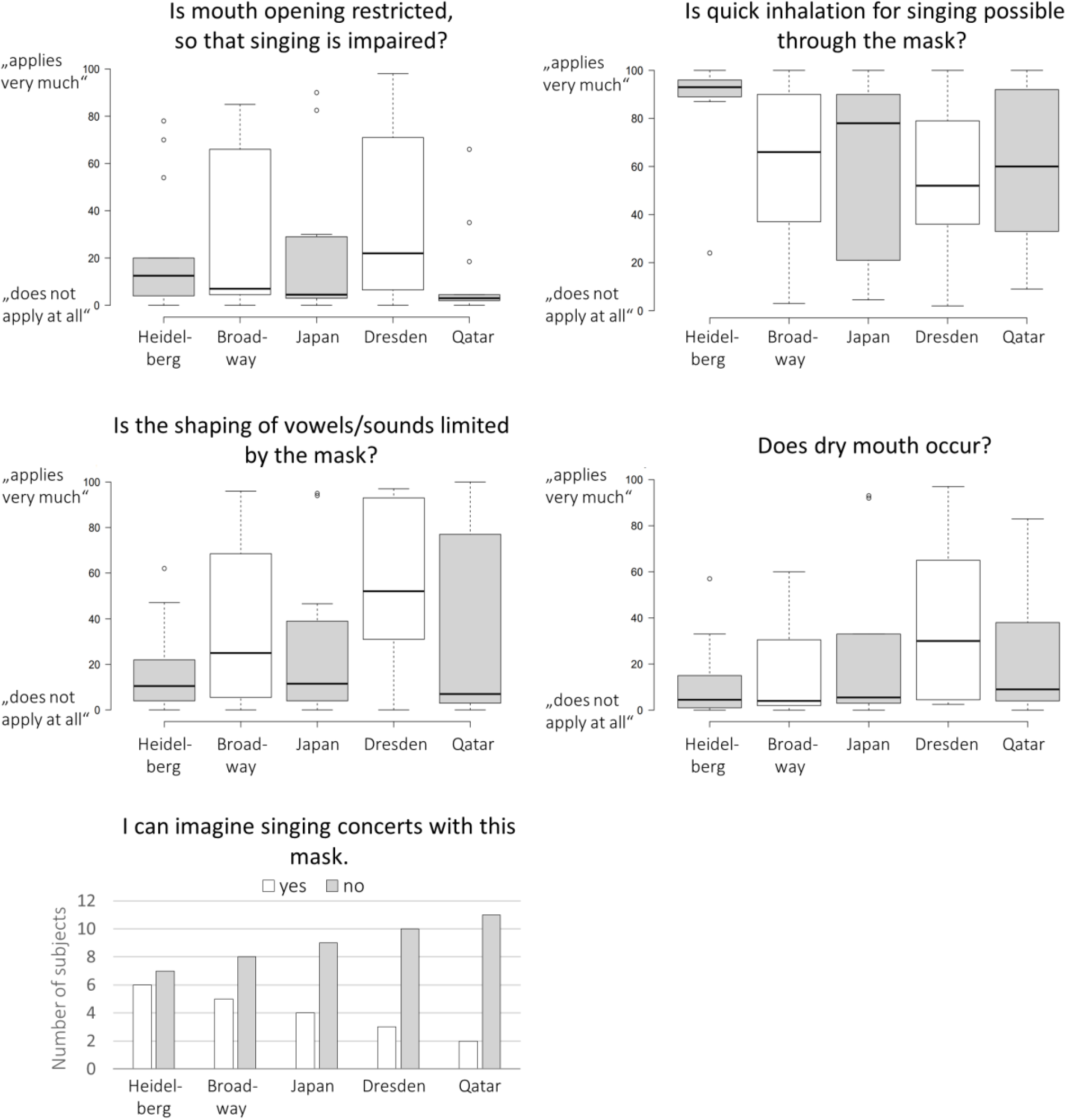
Answers to the different questions of the questionnaire concerning singing without any vapor.

## Discussion

The presented investigation analyzed the impulse dispersion by professional singers wearing different types of singers’ masks, which should reduce the aerosol emission, while still enable the professional singers to perform. In general, it has been found that all the tested masks diminished the impulse dispersion to the front by deflecting the expelled aerosol in sideward and upward direction^40^. Further, most singers’ masks were rated negatively by the professional singers, concluding that a singing performance with the tested masks would not be desirable.

During the CoVID 19 pandemic singing activities have been restricted because of an estimated high transmission risk, which has been concluded from fatal infectious events during choir rehearsals and performances^1-4^. While the aerosol generation is associated with the voice production itself – and therefore cannot be reduced during singing – the transmission pathway could only be blocked or modified at the impulse dispersion, the convectional flow dispersion within closed rooms or by influencing the intake rate of droplets and aerosol particles during inhalation. In the previous study^17^, using the same tasks of melody, pitch and text, the impulse dispersion has been found at a maximum of 1.36 m for singing. Comparable to this, the presented study measured the maximum dispersion for singing without mask at a distance of 1.14 m at the end of the task. It should be mentioned that most of the subjects took part in both studies, which were 8 months apart.

With regard to a blockage of the impulse dispersion of aerosols, it has been shown for many activities, such as breathing and speaking, that surgical masks and FFP-2 masks are able to slow down or stop the dispersion of the impulse ^18,40-42^. Still, professional singing with such conventional masks appeared problematic because of associated dryness of the mouth and impairment of mouth motion, which is more exaggerated and, thus, especially crucial in professional singing. It has additionally been shown that FFP-2 masks reduce intensities in the overtone spectrum up to -12dB at 2000-7000Hz^43^. Furthermore, for a single subject it has been shown that face shield masks do not have much of an impact on the impulse dispersion during singing^18^.

Therefore, many attempts have been made to introduce singers’ masks. The tested masks are only a selection of an internet research, conducted in November 2020, and therefore do not reflect all products and attempts on the market. However, most of the masks had a strong influence on the impulse dispersion to the front, exhibiting lower values for most masks than 1m, with the lowest median dispersion for the Japan Mask. Consequently, the recommended safety distance to the front of 2 - 2.5m ^17^ could be reduced by wearing such masks. At the same time, the dispersion to the side (y-direction) was slightly higher. A small convectional flow, which had already been observed in our previous study^17^, was also present in this study, however, this time by a much smaller amount. Having mentioned this, the distances from the very left to the very right expansion of the expelled vapor cloud when singing with a mask, showed a maximum value of 1.16 m. Therefore, the earlier recommended safety distance to one side of 1.5m could still be justified. Similar to the expansion increase to the sides, the maximum expansion of the cloud in upward (z-)direction was also increased presumably supported by the thermal lift^44,45^. This effect could be beneficial if a ventilation system is assumed with a vertical upward directed net flow with the exhaust openings in the ceiling of a room.

There were many construction differences among the examined masks. Consequently, the evaluation by the singers concerning the practical use of the masks during a performance was very diverse. For most singers, however, the associated limitations concerning inhalation, articulation etc. were so weighty that a singing performance with any of the tested mask models appeared not desirable. A particular problem was the self-auditory perception. In this respect, some masks, especially when surrounding the auricles (Qatar Mask), strongly influenced the perception of the own voice. Therefore, the specific acoustic alteration due to the tested masks should be analyzed in future investigations.

There were many limitations associated with the presented investigation. First, it appears important that the analyzed data only present the primary impulse dispersion by singing and refer to artificially introduced aerosols that do not reflect the real number of potentially virus-laden particles. Consequently, an analysis on the absolute aerosol reduction by the tested masks cannot be offered here. Next, only professional singers have been included. However, a potentially different behavior in amateur singers could not be excluded. Further, it could be speculated that on opera stages the exaggerated articulation could lead to different dispersion patterns, as well as the influence of greater movement activities on the masks and the aerosol clouds. Additionally, the data only refers to a part of a classical masterpiece. Different data when singing other styles or other languages cannot be excluded. Especially Musical Theater singing could lead to observable changes due to its practice of singing with very high pressure. The effects for different singing repertoire and classifications should be analyzed in future investigations. The presented study addresses the aerosol dispersion during a phonation time of about 6 seconds for all tasks. As a result of this rather short time interval in contrast to a realistic rehearsal or performance situations, no possible accumulation effects after a longer time period could be analyzed. However, the chosen time interval was important to allow a better control of the inhaled vapor. Owing to the large number of inhaled aerosols, the mixture possesses a higher density than air. This changed some subjects’ self-awareness during singing – presumably due to changes of the resonance properties of the vocal tract. Because only professional singers were included, all but one subject could, nevertheless, easily compensate for these changes during the experiment. They were able to apply their singing technique under the experimental circumstances. In the experiment, much effort was made to provide the best labor setting possible in order to avoid other influences. However, such influences, particularly the convectional flows within closed rooms, are of great importance. It could be assumed, that convectional flows further distribute the high-concentration-cloud of aerosol among the other singers and even among the audience. Therefore, an active and permanent ventilation is of great importance to sufficiently remove aerosols. In the presented experiment, the investigators tried to use the masks to their best of their understanding. However, after analyzing the Heidelberg-mask the producers provided further information on different usages of the mask. They remarked that an inlay, placed between the two layers of mesh, could potentially reduce the dispersion to the front. Therefore, it cannot be excluded that this kind of modification could influence the data. A further limitation was that, in contrast to the first experiment^17^, no soft versions, no spoken text, nor sung vocalizes without consonants have been analyzed. However, the previous study ^17^had shown, that the greatest dispersion occurred in singing loudly with text, and, consequently, exhibited the potentially highest risk that had to be focused on. Last, because only professional singers were included, the number of subjects was too small to perform comparative statistical analyses. We hope that a large number of singers will be included in future studies in order to check the observed differences.

## Conclusions

It has been shown that singers’ masks influence the impulse dispersion of inhaled artificial aerosols demonstrating lower distances to the front. However, as shown by the answers to the questionnaires improvements are necessary to enable the practical implementations for professional singing.

## Data Availability

Parts of the data (exceot Image material in which the subjects could be recognized) are available on request

## Acknowledgements

The study was supported by the State of Bavaria, Ministry for Science and Arts.

